# Genomic characterization of *Streptococcus pneumoniae* isolates among pediatric patients in Addis Ababa, Ethiopia

**DOI:** 10.1101/2024.06.21.24309271

**Authors:** Abel Abera Negash, Ana Ferreira, Daniel Asrat, Abraham Aseffa, Piet Cools, Leen Van Simaey, Paulina A Hawkins, Lesley McGee, Mario Vaneechoutte, Stephen D Bentley, Stephanie W Lo

**Affiliations:** Armauer Hansen Research Institute (AHRI), Addis Ababa, Ethiopia; Parasites and Microbes, Wellcome Sanger Institute, Hinxton, UK; Department of Microbiology, Immunology and Parasitology, School of Medicine, Addis Ababa University, Addis Ababa, Ethiopia; Laboratory Bacteriology Research, Department of Diagnostic Sciences, Faculty of Medicine and Health Sciences, Ghent University, Belgium; Centers for Disease Control and Prevention, Atlanta, GA, USA

## Abstract

**Background and aims:** Despite the introduction of pneumococcal conjugate vaccines (PCV), *Streptococcus pneumoniae* still remains an important cause of morbidity and mortality, especially among children under 5 years in sub-Saharan Africa. We sought to determine the distribution of lineages and antimicrobial resistance genes of *S. pneumoniae*, 5-6 years after the introduction of PCV10 in Ethiopia.

**Methods:** Whole genome sequencing (WGS) was performed on 103 *S. pneumoniae* (86 from nasopharyngeal swabs, 4 from blood and 13 from middle ear swabs) isolated from children aged < 15 years at three health care facilities in Addis Ababa, Ethiopia from September 2016 to August 2017. Using the WGS data, serotypes were predicted, isolates were assigned to clonal complexes, Global Pneumococcal Sequence Clusters (GPSCs) were inferred and screening for alleles and mutations that confer resistance to antibiotics was performed using multiple bioinformatic pipelines.

**Results:** The 103 *S. pneumoniae* isolates were assigned to 45 different GPSCs. The most common GPSCs were GPSC1 (sequence type (ST) 320, serotype 19A), 14.6%; GPSC268 (ST 6882 and Novel STs; serotypes 16F, 11A and 35A), 8.7% and GPSC10 (STs 2013, 230 and 8804; serotype 19A), 7.7%. Intermediate resistance to penicillin was predicted in 14.6% of the isolates and 27 different Penicillin Binding Protein (PBP) allele combinations were identified. Variations in sulfamethoxazole-trimethoprim (*fol*A and/or *fol*P), tetracycline (*tet*M, *tet*O or *tet*S/M) and macrolide (*erm*B and and/or *mef*A) resistance genes were predicted in 66%, 38.8% 19.4% of the isolates, respectively. Multidrug resistance (≥ 3 antibiotic classes) was observed in 18.4% (19/103) of the isolates and 78.9% of them were GPSC1 (ST320, serotype 19A).

**Conclusion:** Five to six years after introduction of PCV10 in Ethiopia, the population of *S. pneumoniae* is quite diverse, with the most common lineage an MDR GPSC1 (ST 320, Serotype 19A), which is not covered by the PCV10. Continued assessment of the impact of PCV on the population structure of *S. pneumoniae* in Ethiopia is warranted.

**Impact statement:** This study provides a detailed analysis of the genomic features of carriage and disease *Streptococcus pneumoniae* isolates from paediatric patients in Addis Ababa Ethiopia collected 5-6 years after introduction of PCV10 in the country. The study describes the distribution of serotypes, lineages, resistance genes and *in silico* predicted phenotypic antimicrobial resistance. The study highlights the predominance of multidrug resistant serotype 19A expressing GPSC1 (CC320). The findings underline the importance of continued genomic surveillance of pneumococcal carriage and disease to understand the selective pressure of vaccines on lineages and associated antimicrobial resistance.

**Data Summary:** Genome sequences are deposited at ENA with accession numbers (ERR9796440-ERR9990857, ERR10419695-ERR10419739). The authors confirm all supporting data have been provided within the article or through supplementary data files.

## Introduction

*Streptococcus pneumoniae* is an important cause of morbidity and mortality. It caused an estimated 318,000 deaths in 2015 in children aged 1-59 months globally, and more than 50% of these deaths were in the African continent. This is despite the introduction and use of pneumococcal conjugate vaccines (PCV) between the years 2000-2015, which already caused a 50% reduction in pneumococcal deaths [1].

Currently, over 100 pneumococcal serotypes have been identified [2], while the available conjugate vaccines target only a limited number of the most prevalent serotypes causing invasive disease. Introduction of PCV in the routine immunization of infants has proved to be effective in reducing vaccine type (VT) carriage [3] and the burden of pneumococcal diseases caused by VT pneumococci [4]. In addition, indirect benefit of PCV against VT colonization and disease in the unvaccinated population (herd protection) has also been documented [5]. The overall pneumococcal carriage prevalence has however stayed the same mainly due to a significant increase in non-vaccine type (NVT) pneumococci, indicating that the NVTs are occupying the vacant niche. Likewise, an increase in pneumococcal disease due to NVT was also observed in many European countries and the US after introduction of PCV7 [6]. A similar trend has been reported from both European and African countries after introduction of PCV10 and PCV13 [7,8][9].

Whole genome sequencing (WGS) has become a valuable tool for understanding the distribution of pneumococcal lineages, population evolution and serotype switching and replacement [9] [10]. The international definition of pneumococcal population structure known as global pneumococcal sequence clusters (GPSC) was defined to serve as a unifying tool for epidemiological analysis of pneumococcal populations in the post-PCV era [11].

In a recent study, using WGS, Corcoran *et al.* [10] were able to identify sub-clade/variants of serotype 19A associated with invasive pneumococcal disease (IPD) in vaccinated children in Ireland, indicating vaccine failures after PCV13 introduction [12] and highlighting the importance of WGS in studying the molecular epidemiology of *S. pneumoniae*.

In Ethiopia, the PCV10 GSK Synflorix® (serotypes 1, 4, 5, 6B, 7F, 9V, 14, 18C, 19F and 23F) was introduced in October 2011 as a three-dose primary series without a booster dose (3p+0). This was replaced with a PCV13, Pfizer Prev(e)nar® (PCV10 serotypes plus serotypes 3, 6A, and 19A), in July 2020 and is currently the only PCV used.

The aim of the study was to determine the distribution of serotypes, lineages, and antibiotic resistance determinant genes among *S. pneumoniae* isolates collected from pediatric patients in Addis Ababa, Ethiopia 5-6 years after the introduction of PCV10.

## Methods

### Bacterial isolates and serotyping

The *S. pneumoniae* isolates included in the present study were isolated from children aged 0–15 years attending pediatric emergency departments in four major health care facilities in Addis Ababa, Ethiopia (Tikur Anbessa Hospital, Yekatit 12 Hospital, Girum Hospital and Dr. Yared Pediatric Specialty Center) from September 2016 to August 2017. *S. pneumoniae* was isolated from nasopharyngeal swabs of children with community acquired pneumonia (n = 73), nasopharyngeal swabs of children with non-respiratory illnesses (n = 13), blood of children with pneumonia (n = 1) or sepsis (n = 3) and middle ear swabs of children with acute otitis media (n = 13) and their serotypes which were determined by Quellung reaction [13] using antisera, obtained from the Statens Serum Institut (Copenhagen, Denmark) have been reported in previous studies [14–17].

### Genomic characterization

Genomic DNA was extracted using PureLink™ Genomic DNA Kit (Invitrogen, Carlsbad, CA). The Illumina HiSeq platform was used to produce paired-end reads with a length of approximately 150 base pairs. Assembly of sequences was performed using the Sanger assembly pipeline velvet [18] and sequences were mapped to the genome sequence PMEN1 of strain ATCC 700669. Quality of the sequences was assessed using R script (https://github.com/StephanieWLo/Genomic-Surveillance). Quality score parameters used for selection were: sequencing depth of > 20X, > 60% map to reference genome PMEN1, assembly length of 1.9 Mb - 2.3 Mb, less than 500 contigs and less than 220 heterozygous SNP sites. The assembled reads were annotated using Prokka (https://github.com/tseemann/prokka) [19]. Serotypes were confirmed using SeroBA (https://github.com/sanger-pathogens/seroba) [20]. WGS based multi-locus sequence typing was performed as previously described [21] and isolates were assigned to sequence types (STs) and clonal complex (CC) using a single locus variant threshold [22], and clustering the sequences into lineages and Global Pneumococcal Sequence Clusters (GPSCs) was inferred using PopPUNK (https://poppunk.net/) [23]. Screening for alleles and mutations that confer resistance to antibiotics was performed using ARIBA [24]. Penicillin-binding protein transpeptidase amino acid sequence types (PBP types), which were based on pbp1A, pbp2B, pbp2X changes and penicillin minimal inhibitory concentrations (MIC), were determined as previously described [25][26]. Phenotypic resistance and genes for chloramphenicol (cat), cotrimoxazole (folA and folP), erythromycin (ermB and mefA), tetracycline (tetM, tetO and tetS/M), and vancomycin (vanA, vanB, vanC, vanD, vanE and vanG) were predicted from genomic data, as previously described [27–29]. Multidrug resistance was defined as resistance to three or more classes of antibiotics. Phylogenetic analysis was performed by constructing a maximum-likelihood tree using FastTree [30]. Lineage specific phylogenetic analysis was performed using Gubbins (<u>https:// github.com/sanger-pathogens/Gubbins</u>).

### Ethical considerations

The study procedures were in accordance with the Helsinki Declaration. The study protocol was approved by the AHRI/ All Africa Leprosy Rehabilitation and Training Hospital (ALERT) Ethical Review Committee (AAERC) (PO/017/15) and the National Research Ethics Review Committee (No. 310/194/17). Official permission letter was obtained from all the study sites. A written informed assent and consent was obtained from study participants, and parents or guardians of children respectively, before including them in the study. The study participant’s right to refuse or not give samples without affecting their routine medical services was granted. Samples were coded to keep the confidentiality of the study participants’ personal information.

## Results

### Prevalence of pneumococcal serotypes

The concordance between Quellung serotypes and the predicted serotypes on the basis of WGS at serotype level was 77.7% for the 103 isolates, while the concordance at serogroup level was 82.5% (Supplementary table 1). A total of 36 and 38 different serotypes were identified by Quellung and SeroBA, respectively. The most common serotype was serotype 19A (28.4%), followed by 16F (7.8%). Serotype 19A was the most common serotype in all the three sample types (nasopharyngeal swabs, blood, and middle ear swabs). Among the serotypes identified, only 5.8% were PCV10 (GSK) serotypes (*i.e.*, serotypes 6B (n=1), 7F (n=1), 19F (n=3) and 23F (n=1)). PCV13 (Pfizer) would increase the coverage from 5.8% to 38.8%. The overall coverage for PCV10 (SII), PCV15 (Merck), PCV20 (Pfizer), PCV24 (Merck) and IVT-25 (Inventprise) were 38.8%, 38.8%, 48.5%, 48.5% and 51.5%, respectively (Supplementary table 2).

### Pneumococcal lineages

The 103 *S. pneumoniae* isolates were assigned to 46 different GPSCs (Figure 1, Supplementary table 2), of which 40 were already present in the GPSC reference database (www.pneumogen.net\\gps\\assigningGPSCs.html). GPSCs # 1004, 1005, 1013, 1014, 1015, and 1016 were assigned to the novel lineages. The most common GPSCs were GPSC1 (sequence type (ST) 320, serotype 19A), 14.6%; GPSC268 (ST 6882 and Novel STs; serotypes 16F, 11A and 35A), 8.7%) and GPSC10 (STs 2013, 230 and 8804; serotype 19A), 7.7% (Table 1). These all belong to the most common globally circulating lineages. GPSC1 was the most common lineage in isolates from both nasopharyngeal swabs (8/87) and middle ear swabs (6/13). The 103 isolates sequenced were assigned into 73 known STs and 30 novel STs. Among known STs the most common was ST320. There were three lineages (GPSC1, GPSC10 and GPSC5) that expressed serotype 19A.

**Table 1.**
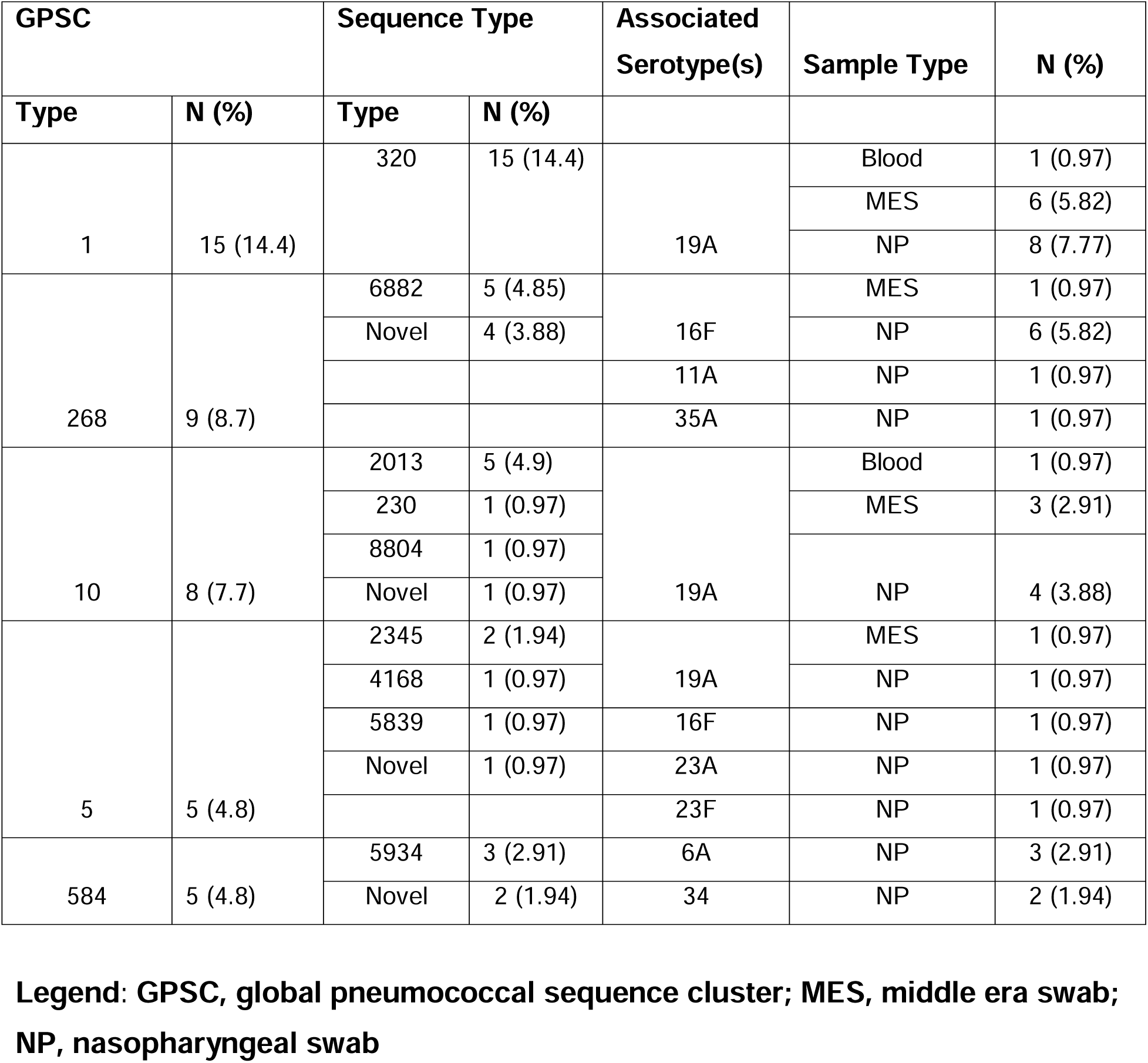
Top five global pneumococcal sequence clusters (GPSCs), associated serotypes and sample sources among pediatric patients in Addis Ababa, Ethiopia.

| GPSC |  | Sequence Type |  | Associated Serotype(s) | Sample Type | N (%) |
| --- | --- | --- | --- | --- | --- | --- |
| Type | N (%) | Type | N (%) |  |  |  |
| 1 | 15 (14.4) | 320 | 15 (14.4) | 19A | Blood | 1 (0.97) |
|  |  |  |  |  | MES | 6 (5.82) |
|  |  |  |  |  | NP | 8 (7.77) |
| 268 | 9 (8.7) | 6882 | 5 (4.85) | 16F | MES | 1 (0.97) |
|  |  | Novel | 4 (3.88) |  | NP | 6 (5.82) |
|  |  |  |  | 11A | NP | 1 (0.97) |
|  |  |  |  | 35A | NP | 1 (0.97) |
| 10 | 8 (7.7) | 2013 | 5 (4.9) | 19A | Blood | 1 (0.97) |
|  |  | 230 | 1 (0.97) |  | MES | 3 (2.91) |
|  |  | 8804 | 1 (0.97) |  | NP | 4 (3.88) |
|  |  | Novel | 1 (0.97) |  |  |  |
| 5 | 5 (4.8) | 2345 | 2 (1.94) | 19A | MES | 1 (0.97) |
|  |  | 4168 | 1 (0.97) |  | NP | 1 (0.97) |
|  |  | 5839 | 1 (0.97) | 16F | NP | 1 (0.97) |
|  |  | Novel | 1 (0.97) | 23A | NP | 1 (0.97) |
|  |  |  |  | 23F | NP | 1 (0.97) |
| 584 | 5 (4.8) | 5934 | 3 (2.91) | 6A | NP | 3 (2.91) |
|  |  | Novel | 2 (1.94) | 34 | NP | 2 (1.94) |

**Legend: GPSC, global pneumococcal sequence cluster; MES, middle era swab; NP, nasopharyngeal swab**

**Figure 1.**
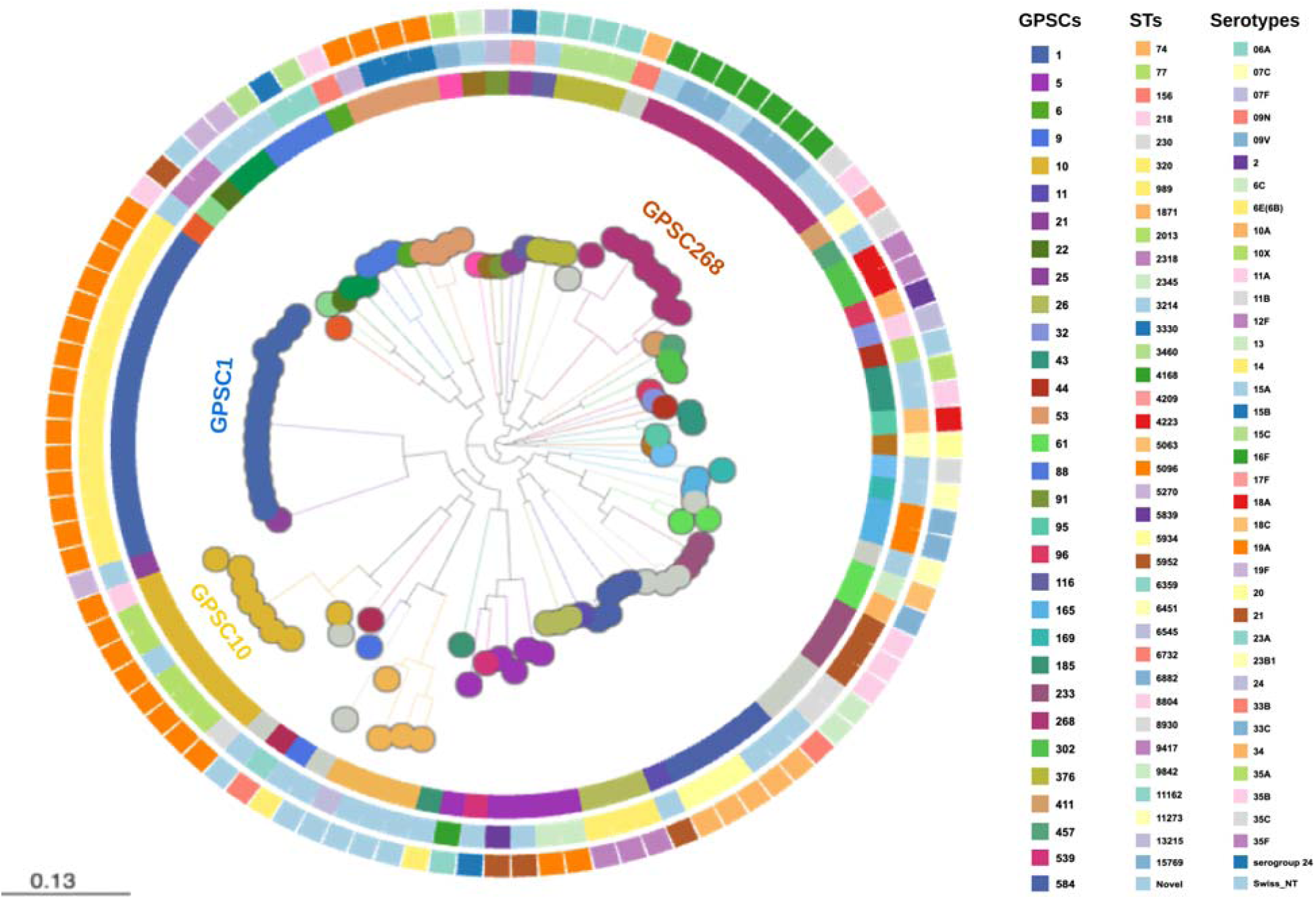
Phylogenetic map indicating the lineages (GPSCs), STs and serotypes for 103 *Streptococcus pneumoniae* isolates from pediatric patients in Addis Ababa, Ethiopia. The inner circle represents GPSCs, the middle circle represents the associated STs, and the outer circle represents the serotypes.

### *In silico* predicted phenotypic antimicrobial susceptibility pattern

The highest level of predicted resistance was seen against trimethoprim-sulfamethoxazole, followed by tetracycline and doxycycline (Figure 2). Intermediate non-susceptibility (MIC 4 μg/mL) was predicted for penicillin in 14.6% of the isolates and for trimethoprim-sulfamethoxazole (MIC 1/19-2/38) in 9% of the isolates. All the intermediate non-susceptible penicillin isolates were serotype 19A. Multidrug resistance (MDR) was observed in 21.3% of the 103 isolates. The majority (72%) of MDR isolates were serotype 19A, expressing GPSC1 lineage (ST 320 (15/16) and ST 230 (1/16)).

**Figure 2.**
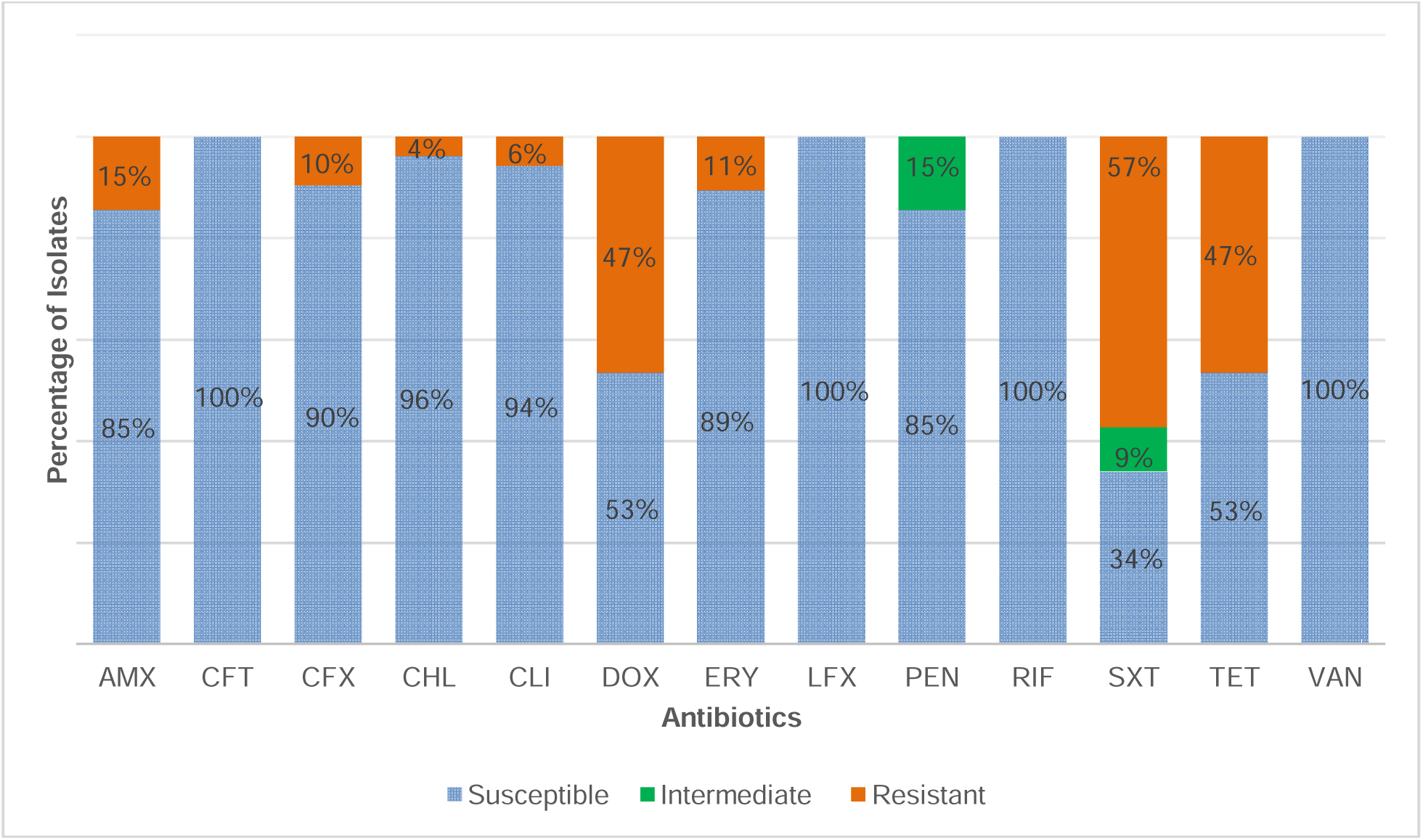
In silico predicted antimicrobial susceptibility pattern of *Streptococcus pneumoniae* isolates among pediatric patients in Addis Ababa, Ethiopia. AMX, Amoxicillin; CFT, cefotaxime; CFX, Cefuroxime; CHL, Chloramphenicol; CLI, clindamycin; DOX, doxycycline; ERY, Erythromycin; LFX, levofloxacin; PEN, Penicillin; RIF, rifampin; SXT, trimethoprim-sulfamethoxazole; TET, tetracycline; VAN, vancomycin.

### Antimicrobial resistance genes

Among isolates with predicted intermediate resistance to penicillin, 27 different PBP combinations were identified; the most frequent PBP combination was 13-11-16, 14.6%(15/103) followed by 90-1-169, 5.8% (6/103).

Changes in *fol*A and/or *fol*P were identified among 61.2% of the isolates, while tetracycline (*tet*M, *tet*O or *tet*S/M) and macrolide (*erm*B and *mef*A) resistance genes were found in 38.8%, 18.4%, and 17.6% of the isolates, respectively (Figure 3). Multidrug resistance (MDR) was predicted in 56.3% of the isolates and the most common MDR lineage was GPSC1 (ST 320, Serotype 19A) (Figure 4).

**Figure 3.**
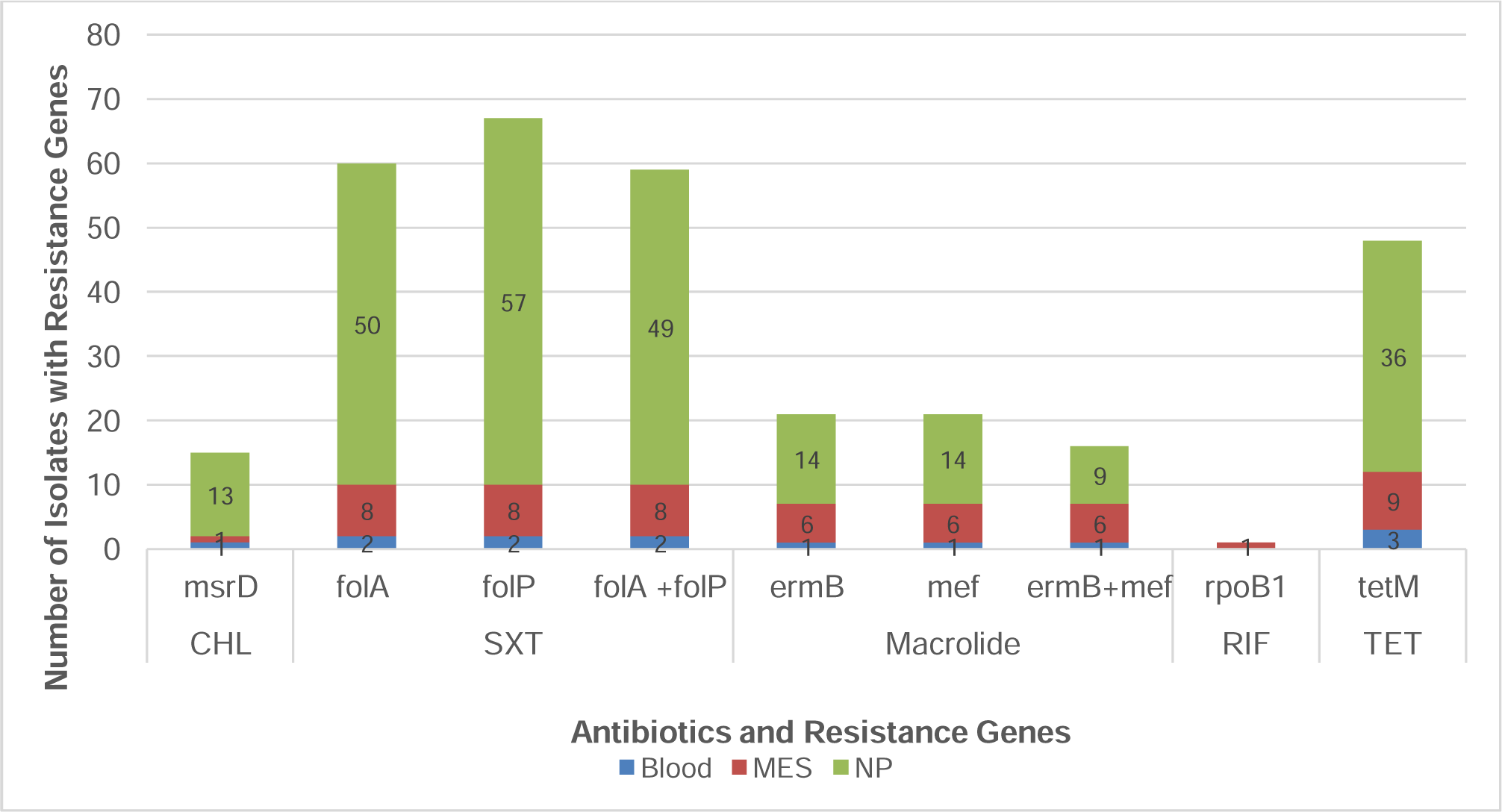
Distribution of non-beta lactam antimicrobial resistance genes among *Streptococcus pneumoniae* isolates from pediatric patients in Addis Ababa, Ethiopia. (CHL, Chloramphenicol; SXT, trimethoprim-sulfamethoxazole; RIF, rifampin; TET, tetracycline; MES, middle ear swab; NP, nasopharyngeal swab)

**Figure 4.**
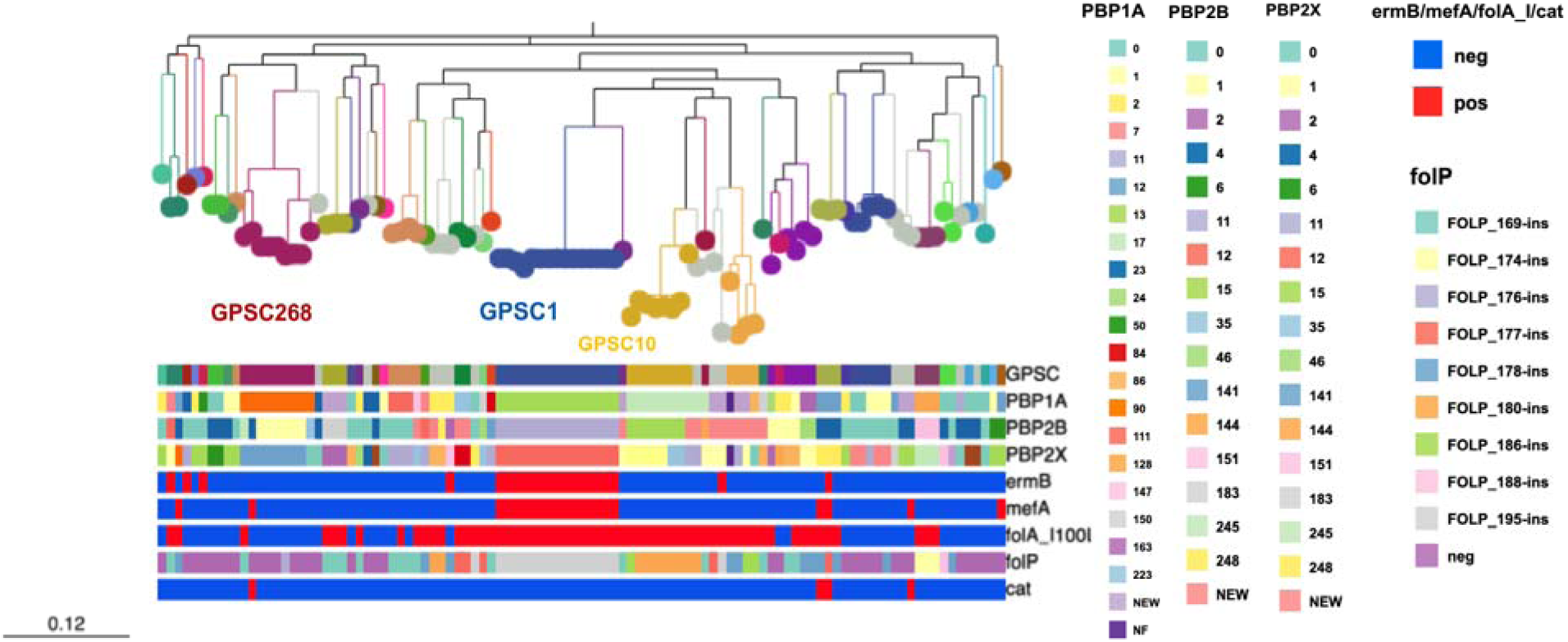
Distribution of resistance genes within different lineages (GPSCs) of *Streptococcus pneumoniae* among paediatric patients from Addis Ababa, Ethiopia. (PBP: Penicillin-binding protein; erm: erythromycin ribosome methylase; mef: macrolide efflux; fol, dihydrofolate reductase; cat, chloramphenicol acetyltransferase)

## Discussion

In this study, we analyzed the genomic characteristics of 103 *Streptococcus pneumoniae* isolates from children with pneumonia (73 isolates from nasopharyngeal swabs and 1 isolate from blood), non-respiratory illnesses (13 isolates from nasopharyngeal swabs), sepsis (3 isolates from blood) and acute otitis media (13 isolates from middle ear swabs) in Addis Ababa, Ethiopia, 5-6 years after introduction of PCV10 in the country. The predominant serotype was serotype 19A (28.4%) followed by 16F (7.8%). In Mozambique, a 40% increase in the colonization prevalence for the three PCV13-specific serotypes (3, 6A, 19A) was reported three years after introducing PCV10[31]. In Nepal and Pakistan, a significant increase in the prevalence of serotype 19A has been reported 4 and 5 years after introduction of PCV10 respectively [32,33]. In Belgium, 4 years after a shift from PCV13 to PCV10, the prevalence of serotype 19A carriage among children in day care centers doubled and became the most dominant serotype [34]. Comparable results were reported in a study from Slovakia, where serotype 19A was the predominant serotype causing acute otitis media among children after the introduction of PCV10 [35]. In Brazil, which introduced PCV10 in 2010, serotype 19A was the leading serotype in all ages from 2014 to 2021 and was responsible for 28.2%-44.6% of all IPD cases in children under 5 years [36]. Since pre-PCV10 serotype data in Ethiopia are scarce, and the association between the prevalence of serotype 19A and the introduction of PCV10 cannot be determined, it will be important to continue to study the epidemiology of serotype 19A and the impact of PCV13, which has replaced PCV10 in the immunization schedule.

The most common pneumococcal lineage in this study was GPSC1, followed by GPSC268 and GPSC10. In Brazil, in the post-PCV10 era, GPSC1 was the third most common lineage causing IPD across all ages and the most common one among children aged < 5 years [37]. Genomic characterization of *S. pneumoniae* isolates from cerebrospinal fluid, nasopharyngeal swabs and non-sterile site swabs in Russia also indicated that GPSC1 was the most common lineage [38]. In a study that investigated pneumococcal carriage dynamics before and after antibiotic treatment in 965 unvaccinated infants and a subset of their mothers in Thailand, GPSC1 was the most common lineage detected after treatment [39].

Two major mechanisms for the evolution of pneumococcal population to vaccine-induced pressure have been suggested. One is through the expansion of lineages that can express only non-vaccine serotypes while the second is the selection of a non-vaccine type serotype expressing lineages among lineages that express both vaccine and non-vaccine serotypes [40]. GPSC1 is a very diverse lineage and consists of more than 28 serotypes and more than 132 STs (https://www.pneumogen.net/gps/GPSC-ST.html). The most common serotypes included in this lineage are serotypes 3, 6C, 14, 19A, 19F and 23F [11]. Since PCV10 has now been replaced with PCV13 in Ethiopia, it will be important to investigate the evolution of GPSC1 expressing serotype 19A.

The acquisition of resistance genes is one of the major strategies by which pneumococci and other pathogens respond to the use of antibiotics and ensure the persistence of lineages[41][37]. In our study, the highest level of predicted resistance among *S. pneumoniae* isolates was seen against trimethoprim-sulfamethoxazole followed by tetracycline and doxycycline. We also identified resistance determinants that confer resistance to chloramphenicol, rifampicin, and tetracycline. Concurrently, the most common resistance mechanism identified regarded changes in *fol*A and *fol*P that confer resistance to sulfamethoxazole-trimethoprim. In Pakistan, 89% of the nasopharyngeal isolates collected 2-6 years after the introduction of PCV10 were resistant to sulfamethoxazole-trimethoprim [42]. Similarly, in Brazil, there was a significant level of resistance to sulfamethoxazole-trimethoprim with changes in *fol*A/*fol*P, among *S. pneumoniae* isolates causing IPD, despite a decline in resistance in the post-PCV10 period (38%) as compared to the pre-PCV10 period (57%), [37]. There were no penicillin-resistant *S. pneumoniae* isolates in this study but intermediate susceptibility to penicillin was seen in 14.6% of the isolates. In Kenya, two years after the introduction of PCV10, there was a significant decline in the carriage of PCV10-type penicillin-intermediate *S. pneumoniae* isolates which made up more than 99% of the penicillin non-susceptible isolates in the pre-PCV10 period [43].

The impact of vaccines on antibiotic resistance is twofold. Not only do they result in direct reduction of strains that carry antibiotic resistant genes, since resistance is often higher among strains that are targets of vaccines, but they also reduce the frequency of febrile infections and therefore lead to a reduction in antibiotic use [44].

In the present study, all the intermediate penicillin non-susceptible and most of the multidrug resistant isolates were serotype 19A expressing GPSC1 (ST320) isolates. Serotype 19A and 16F are among the serotypes that frequently colonize the nasopharynx and among the high-ranking serotypes with acquired *pbp* fragments. Among all pneumococcal lineages, GPSC10, GPSC59, GPSC9, GPSC5 and GPSC1 are the top five ranking lineages known to acquire *pbp* through horizontal gene transfer [41]. In a study performed in Russia, GPSC1 was among the four most common lineages that accounted for 65% of MDR isolates [38]. In Brazil, the main lineage associated with multidrug resistance in the post-PCV10 period was GPSC1 (ST320, serotype 19A) [37]. The expansion of this lineage is thought to have been facilitated by capsular switch events from serotype 19F in the pre-PCV period to 19A in the post-PCV period and association with MDR in the post-PCV period [45–48]. Introduction of PCV13 in the immunization schedule of several countries has resulted in sharp and sustained declines in serotype 19A carriage and disease [49]. However, a recent report from Ireland of vaccine breakthroughs/failure cases due to serotype 19A (ST320) GPSC1 lineage among vaccinated children aged < 5 years [12] indicates the importance of continued genomic surveillance even after introduction of PCV13.

The main limitation of this study is the lack of sequenced isolates from the pre-PCV10 period that would have helped to assess the changes in the population structure before and after PCV10 introduction. A second limitation is the small number of invasive disease isolates, and non-uniform representation of carriage and disease isolates.

In conclusion, genomic characterization of pediatric *S. pneumoniae* isolates in Addis Ababa, Ethiopia showed the distribution of important lineages and antimicrobial resistance determinants 5-6 years after introduction of PCV10 in Ethiopia. The results indicate that GPSC1 which expresses the non-PCV10, PCV13 unique serotype 19A is predominant in both carriage and disease. This is accompanied by the predominance of MDR ST320. It is therefore important to continue performing genomic surveillance of carriage and disease pneumococcal isolates in Ethiopia to assess the impact of the recently introduced PCV13 on the serotype distribution and population structure of *S. pneumoniae*.

## Data Availability

All data produced in the present work are contained in the manuscript

## Supplementary

**Supplementary table 1.** *Streptococcus pneumoniae* isolates (n = 21) with discordant Quellung and SeroBA serotyping results.

| **Isolate** | **Serotype (Quellung)** | **Serotype (SeroBA)** |
| --- | --- | --- |
| **1** | 1 | 14 |
| **2** | 6A | 34 |
| **3** | 6A | 34 |
| **4** | 9L | 9N |
| **5** | NT | 19A |
| **6** | 12B | 12F |
| **7** | 12F | 33B |
| **8** | 15B | 15C |
| **9** | 15B | 15C |
| **10** | 16F | 21 |
| **11** | 18A | 18C |
| **12** | 19A | 2 |
| **13** | 23F | 21 |
| **14** | 23F | 13 |
| **15** | 24F | 15B |
| **16** | 33A | 33C |
| **17** | 33B | 10X |
| **18** | 33C | 17F |
| **19** | 34 | 9V |
| **20** | 34 | 35B |
| **21** | 35A | 35C |
| **22** | 38 | 35B |

\*All isolates were from nasopharyngeal swabs except No.21 which was from blood

**Supplementary table 2.** GPSCs and serotypes of 103 *Streptococcus pneumoniae* isolates among paediatric patients in Addis Ababa, Ethiopia, isolated between September 2016 and August 2017.

| GPSC | Associated Serotypes | N (%) | Isolate sample source | Number |
| --- | --- | --- | --- | --- |
| 1 | 19A | 15 | Blood | 1 |
|  |  |  | MES | 6 |
|  |  |  | NP | 8 |
| 5 | 19A | 2 | MES | 1 |
|  |  |  | NP | 1 |
|  | 23F | 1 | NP | 1 |
|  | 23A | 1 | NP | 1 |
|  | 16F | 1 | NP | 1 |
| 6 | 11A | 1 | MES | 1 |
| 9 | 14 | 1 | MES | 1 |
| 10 | 19A | 8 | Blood | 1 |
|  |  |  | MES | 3 |
|  |  |  | NP | 4 |
| 11 | 21 | 1 | NP | 1 |
| 21 | 19F | 1 | NP | 1 |
| 22 | 15A | 1 | NP | 1 |
| 25 | 15B | 1 | NP | 1 |
| 26 | 12F | 2 | NP | 2 |
|  | 12B | 1 | NP | 1 |
| 32 | 7F | 1 | NP | 1 |
| 43 | 35A | 1 | NP | 1 |
|  | 11A | 1 | NP | 1 |
| 44 | 15A | 1 | NP | 1 |
| 53 | 19A | 3 | NP | 3 |
|  | NT | 1 | NP | 1 |
| 61 | 18A | 1 | NP | 1 |
|  | 34 | 1 | NP | 1 |
| 88 | 15C | 2 | NP | 2 |
|  | 15B | 1 | NP | 1 |
| 91 | 24 | 1 | NP | 1 |
| 95 | 18A | 1 | NP | 1 |
| 96 | 19A | 1 | NP | 1 |
| 116 | 23A | 1 | NP | 1 |
| 165 | 33C | 2 | Blood | 2 |
| 169 | 23B | 1 | NP | 1 |
| 185 | 6B | 1 | NP | 1 |
| 233 | 35B | 1 | NP | 1 |
|  | 34 | 1 | NP | 1 |
|  | 38 | 1 | NP | 1 |
| 268 | 16F | 7 | MES | 1 |
|  |  |  | NP | 6 |
|  | 11A | 1 | NP | 1 |
|  | 35A | 1 | NP | 1 |
| 302 | 35F | 2 | NP | 2 |
| 376 | 6A | 3 | NP | 3 |
| 411 | 33C | 1 | NP | 1 |
| 457 | 11B | 1 | NP | 1 |
| 539 | 24F | 1 | NP | 1 |
| 584 | 34 | 3 | NP | 3 |
|  | 6A | 2 | NP | 2 |
| 624 | 11D | 1 | NP | 1 |
| 645 | 35B | 1 | NP | 1 |
| 661 | NT | 4 | NP | 4 |
| 667 | 20 | 1 | NP | 1 |
| 690 | 6C | 1 | NP | 1 |
| 699 | 9L | 1 | NP | 1 |
| 867 | 33B | 1 | NP | 1 |
| 879 | 19F | 2 | NP | 2 |
| 883 | 21 | 1 | NP | 1 |
| 883 | 21 | 1 | NP | 1 |
| 1004 | 10A | 1 | NP | 1 |
| 1005 | 13 | 2 | NP | 2 |
| 1013 | 33B | 1 | NP | 1 |
| 1014 | NT | 1 | NP | 1 |
| 1015 | 7C | 1 | NP | 1 |
| 1016 | NT | 1 | NP | 1 |

